# Developing and testing an integrated patient mHealth and provider dashboard application system for type 2 diabetes management among Medicaid-enrolled pregnant individuals based on a user-centered approach: Mixed-methods study

**DOI:** 10.1101/2022.02.07.22270501

**Authors:** Naleef Fareed, Priti Singh, Emma Boettcher, Yiting Wang, Kartik Venkatesh, Robert Strouse

**Affiliations:** CATALYST – The Center for the Advancement of Team Science, Analytics, and Systems Thinking, College of Medicine, The Ohio State University, Institute for Behavioral Medicine Research, 460 Medical Center Drive, Columbus, OH 43210, United States; Department of Biomedical Informatics, College of Medicine, The Ohio State University, 1800 Cannon Drive, 250 Lincoln Tower, Columbus, OH 43210, United States; Department of Research Information Technology, College of Medicine, The Ohio State University, Suite 275, 530 W. Spring St., Columbus, OH 43215, United States; Division of Maternal-Fetal Medicine, Department of Obstetrics and Gynecology, The Ohio State University, 1800 Zollinger Road, Columbus, OH, 43221, United States

**Author notes:** **Corresponding Author:** Naleef Fareed PhD, MBA, 460 Medical Center Drive, Columbus, OH 43210, United States.

**Keywords:** pregnancy, diabetes-management, mobile application, mHealth, dashboard, user-centered, mixed-method

## Abstract

**Background:** Meaningful integration of mobile health (mHealth) applications that capture and provide feedback on important dimensions is lacking and is required to promote behavioral changes that are linked to better maternal and birth outcomes among pregnant individuals. The design and use of digital health technologies among individuals covered by public health insurance is typically overlooked and has implications for how this group can manage their health with the support of technology. Medicaid-enrolled patients with type 2 diabetes (T2D) during pregnancy struggle to manage their diabetes due to clinical and social needs barriers. There is an opportunity to develop a tailored and integrated technology solution (patient mHealth application and provider dashboard) that provides a 360-degree view of the patient for this group that could improve health outcomes and address health inequities.

**Objective:** Our objective is to describe a formative study that developed an integrated patient based mHealth and provider dashboard application system for management among Medicaid-enrolled pregnant patients with T2D. Our goals were to: 1) develop a set of user specifications for the mHealth and dashboard applications; 2) develop prototypes based on user needs; and 3) collect initial impressions of the prototypes to subsequently develop refined tools that are ready for deployment.

**Methods:** Study activities followed a double diamond framework with a participatory design mindset. We first conducted a literature review to articulate the problem. Phase I subsequently involved a qualitative inquiry of the needs faced by patients and their providers and current clinical workflows at our AMC (Discover), and brainstorming activities (Define) to ideate and define the value specifications our mHealth and dashboard applications to our end-users. Phase II involved the design and development of low- and high-fidelity prototypes that incorporated a core set of functions based on our Phase I findings and the theoretical underpinnings of social cognitive theory (Develop). We conducted usability and cognitive tests of our high-fidelity prototypes with potential users to gather feedback about the content, function, and aesthetics of the prototypes (Delivery).

**Results:** We recruited seven patients and seven providers in our study. All participants completed Phase I, and three patients (42%) and four providers (57%) completed Phase II. We identified three themes that exemplified pregnancy experience among Medicaid-enrolled individuals with T2D: *managing exhaustion, adherence to a new regimen, and preparing for pregnancy*. Patients expressed a core set of expected features for an mHealth application: *electronically captured and managed information, access to support systems, use of diabetes technology*, and *help through problem solving and personalized recommendations*. Providers indicated a general set of expectations for a dashboard application, including features such as *dynamic and actionable data, unmet social needs, enhanced patient-provider communication*. We developed “as-is” and “to-be” swim lanes to depict clinical workflows and critical gaps, and we designed high-fidelity prototypes using this information. Participants provided notable feedback to improve the mHealth application (e.g., from a content perspective, patients asked for more details about the achievement of goals) and dashboard (e.g., from a functional perspective, providers suggested to add a checklist for patient completion of educational resources related to T2D during pregnancy). For both applications, participants reported scores for the NASA Task Load Survey (TLX) that were in the 20th percentile of national TLX scores.

**Conclusions:** Digital health tools have the ability to transform health care among Medicaid-enrolled patients with T2D during pregnancy, with the goal of managing their blood glucose levels, which is a precursor to experiencing a successful pregnancy and birth. Distilling patient and provider needs and preferences – and then using that information, along with prior studies and theory, to develop applications – holds great potential in tackling complicated health care issues. The methods described in our study can be used as a template for future design considerations specific to the development of digital health interventions, including those focused on understudied populations.

## Introduction

Type 2 diabetes (T2D) affects nearly 100,000 pregnant individuals in the U.S. every year [1,2]. The prevalence of T2D is expected to double in the next decade, putting a greater number of mothers and infants at risk for diabetes-related complications in pregnancy [3]. In parallel, the prevalence of T2D has doubled among pregnant individuals of low socioeconomic status (SES) [4]. Over 50% of pregnant individuals with T2D are enrolled in Medicaid and are of low SES [5,6].

Diabetes management in pregnancy requires strict glycemic control, which necessitates strict lifestyle adjustments, glucose monitoring, and pharmacotherapy over a relatively short amount of time [7]. Suboptimal glycemic control results in adverse maternal and fetal outcomes, including cesarean delivery, preeclampsia, birth trauma, severe maternal morbidity, NICU admission, neonatal hypoglycemia, fetal growth abnormalities, and stillbirth [8]. Clinical care alone is not sufficient, and social determinants of health (SDoH) can have a significant influence on health outcomes [9]. SDoH are the conditions in which people are born, grow, live, work, and age, and they include factors such as insurance status and SES, which is consistently a strong predictor for diabetes onset and progression [9,10]. SDoH impact the ability of Medicaid-enrolled pregnant individuals with T2D to achieve glycemic control through the absence of facilitators such as reliable transportation to attend visits, access to resources to engage in healthy diet and exercise, and convenient methods to log self-monitored glucose values and adjust insulin dosing [11–18]. Limited education and income may further exacerbate barriers to care for Medicaid-enrolled individuals with T2D, who are unable to engage in activities that achieve glycemic control [19]. When social needs are not met, T2D management may become increasingly difficult [4,20–22]. This is concerning because recent studies have documented higher rates of adverse pregnancy-related outcomes among individuals on Medicaid compared to those with commercial insurance, regardless of diabetes status [11,23]. These findings highlight the need for interventions that are tailored to Medicaid-enrolled pregnant individuals with chronic comorbid conditions, including T2D.

Evidence suggests that current models of maternity care are inconsistent with patient preferences regarding diabetes management and interaction with their care providers [24]. Pervasive technology such as mHealth and Internet of Things (IoT) wearables present several opportunities for fostering pregnancy care management [25]. Nearly 75% of pregnant individuals reported downloading at least one mHealth app, and a majority of them indicated they are likely to use them once per week [26,27]. Over 90% of women of reproductive age are smartphone users, including those on Medicaid. [28] Existing research indicates the willingness of pregnant individuals to use digital tools for health management [29,30].

Stand-alone mHealth applications have demonstrated benefits for diabetes management among non-pregnant individuals, but the potential of mHealth among pregnant individuals has yet to be fully realized [21]. A recent study comparing performances of pregnancy-related diabetes management mHealth applications indicated that existing applications perform well in areas of education and information, but only a few provided comprehensive and evidence-based educational content, tracking tools, and the ability to integrate meaningfully with electronic health record systems [21]. Applications that are comprehensive, personalized, and integrated within care team workflows are likely to be more effective [21], increase patient uptake [31], sustain patient and provider behavior change over time [32], and demonstrate long-term sustainability [31]. Existing diabetes related mHealth applications also fail to capture SDoH and address specific patient needs. Current prenatal and diabetes care delivery for Medicaid enrolled pregnant individuals is fragmented [11]. Furthermore, the current health system lacks a multifaceted intervention that incorporates tools at both the patient and provider ends to facilitate patient-provider communication and provide a 360-degree view of patients’ pregnancy experiences.

A provider-facing, bi-directional dashboard can provide comprehensive diabetes information to care teams, including timely clinical alerts about glycemic control, psychosocial issues, and treatment plans; [33] facilitate team-based provider coaching and feedback [34], including working with the patient’s agenda and recognizing patient beliefs, values, and readiness for change; and help with behavioral modification [35,36]. Outside of pregnancy, such an approach has improved glycemic control [37] and adherence to evidence-based diabetes care [33]. Based on the existing evidence, our study team was motivated to explore a platform that capitalizes on the integration of an mHealth application and dashboard technology to provide a comprehensive, tailored, and team-based solution that addresses clinical management and social needs among Medicaid-pregnant individuals with T2D.

## Objective

Our objective is to describe a formative study that developed an integrated, patient-based mHealth and provider dashboard application system for management among Medicaid-enrolled pregnant individuals with T2D. Our goals were to: 1) develop a set of user specifications for the mHealth and dashboard applications; 2) develop prototypes based on user needs; and 3) collect initial impressions of the prototypes to subsequently develop refined tools that are ready for deployment.

## Methods

### The Double Diamond Framework

Our study activities followed the double diamond framework, which is a user-centered approach widely used in information technology development. Iterative development cycles are utilized to produce user-specified needs and preferences. Figure 1 shows the roadmap of our design framework, starting with a diagnosis phase and followed by four sequential double diamond phases: discover, define, develop, and deliver [38]. Our study utilizes the double diamond process to: a) define problems faced by Medicaid-enrolled individuals and their providers for effective diabetes management during pregnancy, b) develop prototypes of our mHealth and dashboard applications, and c) elicit reactions from users with regard to usability and acceptance.

**Figure 1.**
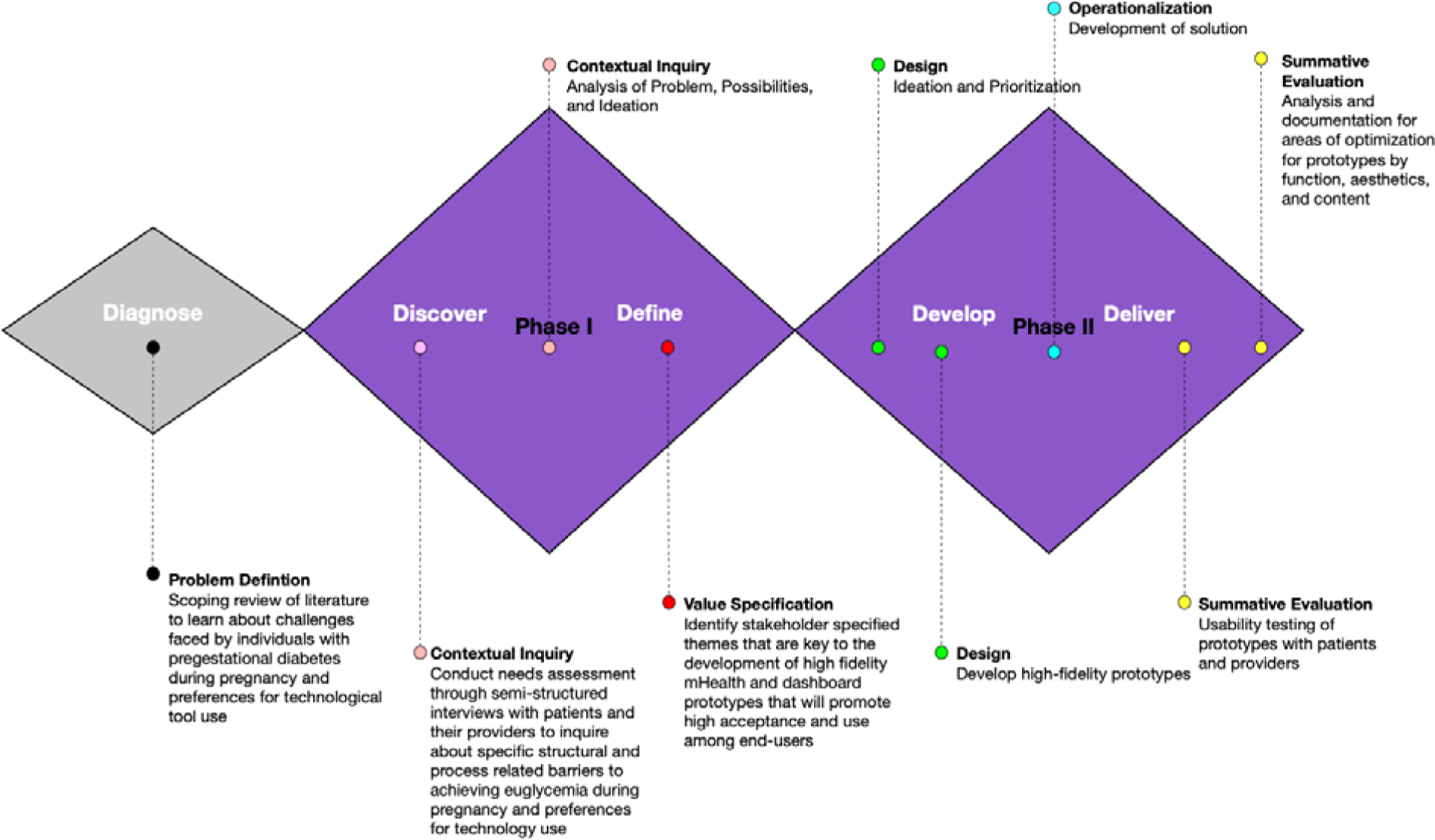
*ACHIEVE* pilot study activities through the lens of a double diamond framework and participatory mindset

The shape of the double diamond figure signifies the process of collecting divergent insights during discovery and subsequently translating those insights into concepts by convergence and definition [39]. This conceptual framework has been successfully used for complex health care problems [40,41]. For example, Asbjørnsen and colleagues designed an eHealth intervention that aimed to support behavior change for long-term weight loss maintenance in the general population [39]. We complemented our double diamond framework thought process with a participatory design mindset and included system requirements generated by end-users to ensure expectations for the final product were met [42].

The section that follows describes each of the double diamond phases as they pertain to our study. Our team used brain mapping exercises aided with a Miro board to outline ideas, collaborate with other team members, identify themes, and design prototypes for the study.

### Diagnose

As a first step, three members of our research team (EB, AL, GO) conducted a scoping review of the literature for key problems and existing gaps related to diabetes management in pregnant individuals. Our research focused on two domains: 1) health disparities due to clinical and non-clinical factors and 2) technology use. Both areas focused on individuals who are at risk, members of minority populations, and on public health insurance. We examined various sources that included peer-reviewed articles and registered clinical trials for scientific evidence. Significant findings were reported on our study Miro board for reference and discussed as a team, which included gaining input from our clinician investigator (KV) for validation from a health care provider perspective.

### Phase I

#### Discover (Formative Interviews)

The problems identified in the diagnosis phase were further studied in our target population by means of a pilot study. Our team conducted individual interviews to identify current structural and process barriers, as well as practices associated with management of diabetes among Medicaid-enrolled patients.

We conducted separate interviews with patients and their providers at our AMC Division of Maternal Fetal Medicine (MFM). Interviews examined three areas of interest: a) current workflows for standard of perinatal and post-partum care; b) current workflows and processes for collection of SDoH-related information to address any unmet social needs, and c) participants’ needs in regard to the design an mHealth application (for patients) or dashboard (for providers) for diabetes management during pregnancy. All study activities were approved by our AMC Institutional Review Board.

### Study Sample

#### Patient Recruitment

Patients were recruited from the Diabetes in Pregnancy Program within the division of Maternal Fetal Medicine. Inclusion criteria included patients who were pregnant, had T2D diabetes, covered by Medicaid insurance, and were able to speak English. Patient information including name and diagnosis were used to identify eligible patient participants. A convenience sample of seven patients was identified, and their contact details were provided by the clinician. Identified eligible patients were recruited and consented by a study team member. Each patient received a $50 gift card at the end of the study (after completing both phases) as an incentive for participation.

#### Provider recruitment

Providers included physicians, nurses, diabetes nurse educators, and nurse assistants who were involved in the care of pregnant patients with diabetes. Providers were recruited from the Diabetes in Pregnancy program. A convenience sample of seven providers was identified, and their professional email addresses were provided by our clinical study team member. Providers were recruited by email and consented by a study team member. Each non-physician provider received a $50 gift card at the end of both study phases as an incentive for participation.

#### Semi-structured Interviews

We interviewed seven patients and seven providers. The interviews typically lasted 60 minutes, were audio recorded, and were transcribed verbatim. Our research team conducted the interviews using semi-structured guides to explore patient and provider experiences. These guides were developed by the research design team, pilot tested on two patients and two providers, and refined based on the provided feedback. The patient interview guide included four key sections: patient background, management of pregestational diabetes, information on social determinants of health (SDoH), and desired features in an mHealth application for diabetes management during pregnancy. Similarly, the provider interview guides included four sections: provider background, treatment of pregestational diabetes, information on SDoH, and desired features in a dashboard for diabetes management during pregnancy.

### Define (Data Analysis)

This phase consisted of thematically organizing patient and provider interviews by means of open coding. Interviews were analyzed inductively to categorize our findings and reveal emergent themes (Fereday & Muir-Cochrane, 2006). Three team members (EB, YW, NF) reviewed at least 20% of each transcript, and two members reviewed (PS, NF) the entire transcripts independently to identified emergent themes. These two members met frequently to ensure consistency of coding and agreement about definitions of new codes as they emerged during the coding process, consistent with the grounded theory approach [43]. A third researcher team member (RS) independently reviewed the thematic codes and reconciled any differences. Information from our literature review and thematic analyses, in concert with the fundamental constructs of social cognitive theory (SCT) [44,45], was used to develop elements that formed high-level specifications for the mHealth and dashboard prototypes. Microsoft Word was used to analyze quotes and STATA 17SE was used to analyze all quantitative data for the study.

### Phase II

#### Develop (Prototype Design and Development)

We designed numerous low-fidelity prototypes in the form of pencil-and-paper sketches to visually explore features based on the key themes identified from the phase I interviews. Many components, features, and interactions represented in the low-fidelity prototypes were then used to inspire the creation of high-fidelity clickable prototypes in Figma for the mHealth and dashboard applications. The features were designed to be user centered, engaging, and driven by the SCT approach to motivate specific behavioral activities among patients to lower their hemoglobin A1c (A1c) through effective management of blood glucose and social needs during pregnancy. We concurrently developed an initial infrastructure architecture to demonstrate integration of the applications via a digital platform with a defined ecosystem within which patients and their providers would engage with the system and communicate with each other.

### Deliver (Usability Assessment)

In the final phase of our pilot study, we used the high-fidelity clickable prototypes of the mHealth and dashboard applications that were developed based on the feedback provided by study participants and our user experience design team. Key features present in the initial low-fidelity prototypes, although not intended to be comprehensive, represented a core set of functions that would promote behavioral modifications as guided by the principles of SCT. Following the approach undertaken by Yardley and colleagues, our research team met multiple times over the course of several weeks to plan, prioritize, and highlight aspects of the prototype features that were crucial [46].

We conducted the Phase II interviews with the corresponding prototypes tested to obtain feedback from the users who participated in our Phase I interviews. We conducted usability testing and think-aloud sessions with patients and providers. The aim of these interviews was to systematically collect feedback about the prototypes using a deductive approach based on our prior work on decision support tools [47,48].We categorized identified themes into one of three categories: (1) function, issues that are related to meaningful and intuitive navigation of an application; (2) content, problems with information provided in an application that complicate or lead to misinterpretation of information; and (3) aesthetics, concerns that impede an application from having a minimalist design that effectively communicates information.

#### Usability and Think-aloud

At the time of interview, each participant was asked to perform a predetermined set of tasks with no time limit. Participants were prompted to complete tasks (e.g., inputting blood glucose values for a specific time of day, setting up a new appointment, and reviewing a goals card). Participant actions and spoken-aloud thoughts during the usability session were recorded for further analysis. Participants were asked to share their experiences about the application and usability of the product. Our team used Zoom to record the session. Our team documented challenges to the use of prototypes and completion of prespecified tasks as a list of recommendations by function, category, or aesthetic.

#### Cognitive Workload Survey

Upon the completion of the usability assessment, participants were asked to respond through Qualtrics to cognitive workload survey questions assessing mental, physical, and temporal demands; frustration levels; overall performance; and effort required to complete the usability assessment. The questions in the survey involved the items from the validated TLX [49].

Feedback from the survey and interviews will be used to develop the final version of the mHealth and dashboard applications.

## Results

### Patient Characteristics

We recruited seven patients and seven providers in our study. All patients and providers completed Phase I, and three patients (42%) and four providers (57%) completed Phase II. Characteristics of our study samples are provided in Tables 1 and 2. As presented in Table 1, most of the patients identified as non-Hispanic Black, with a mean age of 29. Most of our patients had two previous pregnancies (gravida). Among our provider sample, we recruited four physicians and three registered nurses.

**Table 1.** Characteristics of patient sample (n=7)

|  |  |
| --- | --- |
| Age in years<br>(Mean, SD) | 29 ($\pm 3$) |
| Race (n, %) |  |
| Non-Hispanic White | 3 (43%) |
| Non-Hispanic Black | 4 (57%) |
| Gravida (Mode) | 1 (n=2);<br>2 (n=4);<br>3 (n=1) |

**Table 2.** Characteristics of provider sample (n=7)

|  |  |
| --- | --- |
| Gender (n, %) |  |
| Males | 4 (57%) |
| Females | 3 (43%) |
| Role (n, %) |  |
| Physician | 4 (57%) |
| Registered Nurse/<br>Diabetes Nurse Educator | 3 (43%) |

### Diagnosis phase

#### Problem

Pregnant individuals on public insurance with T2D are 50–80% more likely to have elevated A1c levels and experience adverse birth outcomes, which can result from dysglycemia early in pregnancy, compared to their counterparts with private insurance [11,50]. Over 40% of U.S. pregnant individuals receive prenatal care through public insurance [51]. While early and sustained prenatal care improves maternal and neonatal outcomes for T2D [52,53], pregnant individuals on public insurance encounter numerous barriers that preclude appropriate and timely prenatal care [14,54]. Based on the Determinants of Health Model, clinical care alone for this population is not sufficient, and SDoH influence health outcomes [16]. Social needs such as food security, adequate housing, a safe environment, and access to medications and health care influence health outcomes and glycemic control [16]. When social needs are not met, T2D management during pregnancy, including achieving glycemic control, becomes increasingly difficult [9,20].

#### Intervention

Our team proposed an intervention to address social needs among Medicaid-enrolled individuals with poorly controlled T2D during pregnancy. Pregnant individuals are interested in engaging in alternative prenatal care models [55] that can mitigate health and social disparities [54]. In addition, a linked provider dashboard can facilitate better decision making, promote regular contact between patients and providers, and improve patient outcomes [56]. The value of mHealth applications for health-based interventions in diabetes outside of pregnancy has been demonstrated [37,57]. Mobile health applications result in lower A1c levels, improved health care utilization, better patient-reported outcomes (PROs) [58–61], and improved post-intervention engagement and diabetes outcomes. [62] Due to the underlying health inequities and the willingness of our study pregnant population to adopt digital health technology, our team proposed to use mHealth and provider dashboard applications to mitigate existing disparities [29]. In this study, we provide the rationale and process used to develop our initial prototypes.

### Phase I

#### Discover

When asked about their pregnancy experiences, patients indicated common feelings such as constant feeling of tiredness, adherence to a new regimen, and preparing for the pregnancy. Their pregnancy journeys included several lifestyle modifications, barriers, and experiences, and these experiences were captured by our interviews.

#### Managing Exhaustion

When describing their pregnancy experiences, patients frequently reported feelings of constant exhaustion, especially those with older children or employment. Patient 1 explained: “*I’m really tired all the time, I’m so exhausted and then my other child is only two,”* while Patient 2 shared: *“So, we’re always staying active, just because young ones keep you busy, but once I come home, I always feel so exhausted, and I try to like sometimes force myself to go for a walk*.*”* Patient 4 noted: “*Well, definitely sugar, of course, diabetes, feeling very exhausted a lot*.”

#### Adherence to a New Regimen

Diabetes management during pregnancy requires strict medication adherence, frequent assessment of finger stick blood glucose (FSBG) levels upwards of four times per day, and dietary modifications. Patients reported having to be more aware of their health status and willing to adapt to new health regimens, including greater reporting of blood glucose levels, food intake, and physical activities. However, these added measures were perceived as demanding, as indicated by Patient 1: *“I’m taking my medicine better. Before I really wasn’t taking my insulin like that, and now I’m really taking it, taking it better, checking my sugars way more like they asked me to. It’s really tedious, oh my gosh, it’s so tedious*.” This patient also noted that they had to “*just count them [carbs] and make sure that I’m taking the appropriate amount of insulin with it, so it takes a lot of concentration on my side because now it’s like everything is a numbers game with the diabetes*.” Patient 4 shared: *“Basically, I just write everything down that’s how I keep track of everything*.*”*

#### Preparing for Pregnancy

Patients reported the need to prepare for their pregnancy, including with regard to clinical and social needs. Patients recognized having support would be critical to maternal and infant care, especially during the early stages of infant life, as well as the need for financial security. These factors are highlighted in experience shared by Patient 5: *“I feel like I’m more aware of things, like I’m gonna have to take care of somebody else and making sure I’ve got time, money and resources to do that*.*”* They also mentioned: *“I try to plan ahead more than I did. I was, before you called, I was sitting down and writing out a budget, which I’ve never really had to do before. Just to make sure that I would have enough*.*”* Moreover, diabetes management requires significant lifestyle alterations, as reflected in the response of Patient 4: *“It is definitely life changing … with this pregnancy, I was immediately on insulin the very first week I found out. So from there you got to flip life over upside down and basically detox your body from everything you’re used to*.*”* Most patients mentioned that during their prenatal visits, their social needs were not actively discussed in detail. Some mothers requested to be enrolled in the Special Supplemental Nutrition Program for Women, Infants, and Children, and their providers gave them pamphlets and general guidance.

### Patient-reported Needs and Preferences in an mHealth Application

Patient expressed a core set of expectations for an mHealth application: features that electronically captured and managed information, access to support systems, use of diabetes technology, and help through problem solving and personalized recommendations.

### Electronic Capture and Management of Information

Our results indicated the emerging need to include an electronic feature to capture blood glucose levels. Existing processes called for recording blood glucose levels in paper logs and sending them to providers as emails or by fax, which was perceived as being cumbersome, as indicated by Patient 4: “*something that can keep track of my, my sugar records. That would be nice, instead of having to write everything down and then send it on through email. It would be awesome just to have it on the app. That would definitely be a plus*.*”* Patient 1 similarly noted: *“Hey it’s time to take my sugar, let me go ahead and put it in my app instead of writing it on the paper. So we have kids, so the other kids get the paper or you could forget it, stuff like that, but our phones are always in our hands. So that would just make it easier, you know, than that paper. When they showed me the paper, I was like, oh God, I am not going to keep up with it, sorry, no. So that just replacing that would be helpful in general*.*”*

In addition to capturing FSBG values, patients expressed the need to include a feature that captured details about their dietary intake and out-of-range values with means for providing open-ended responses. For example, Patient 4 mentioned: “*it’d be nice to have like a schedule of like your, when you’re supposed to check your sugars and then being able to write down what your sugar was that day and then even going to the extent of what did you eat and explain why your sugar is that low or that high. I think that would be awesome*.*”*

### Support System

Patients suggested the need to include a feature that allows them to communicate with other individuals for support and to learn from their experiences. Patient 3 noted: *“Maybe getting them connected with the right people on how to manage their care. Ways of controlling since they would know*.*”* Patient 4 said: “*What they eat [support group] is a good one because I get bored with the same old food so it’d be nice to know what other people are eating that’s been able to maintain their sugar. What they do, as far as exercise to where they don’t overdo it, but I know walking is a good one, but that would actually be something nice to know what other people is doing to maintain their weight and everything during, especially having diabetes during pregnancy*.*”* They also said: *“Well, it would definitely be nice to have a support system. Like something that I can go to if I need support, especially somebody that already knows what I am going through and how I am feeling. It’s nice to have somebody that can relate*.*”*

### Diabetes Technology

The availability of innovative technology for diabetes management has been rapidly increasing. Patients expressed interest in the use of such technologies, especially continuous glucose monitoring (CGM) devices. Patient 4 expressed interest in using CGM: *“I do think one idea would be awesome would be having a monitor, which I know they have them out there, but for pregnant women … that is actually you’re able to check it like it’s inserted in you. I forget what it’s called, but I know they have some on that out there, but it’d be awesome if they can actually get one approved for pregnant women, to make it so much easier on them*.*”*

Patient 1, who actively used CGM, indicated: *“The scanner that I use is on my phone. I don’t have to have like an additional piece of equipment … I downloaded the app and just scan my arm from there, I don’t have to, you don’t have an extra piece of equipment, so it just makes it easier for them to get the numbers from directly from the app*.*”* This patient also explained the motivation behind switching to CGM: “*My PCP told me about it ’cause they were having so much trouble getting me the finger stick. It just hurts. Like, I won’t lie to you. I’d rather take the insulin than do the finger stick, it, it hurts that bad … even though my insurance doesn’t cover it, I still pay out of pocket for it, because it’s so much easier, I can just stick myself the one time, then I just scan my phone on my arm, way easier. As much as they want me to check my sugar, like I, I am not going to be able to stick myself. It’s something about sticking myself that hurts because you can’t see the needle and … it’s like a jack-in-the-box almost, you’re turning it, you’re turning it, you know it’s going to pop out, when is it going to pop out?”*

### Problem Solving and Personalized Recommendations

Pregnancy is a unique experience for every patient and requires guidance that accounts for specific challenges faced by a pregnant individual. Patients reported the need for direction on how to actively plan activities and meals that aligned with effective blood glucose management. Patient 3 indicated that “*Like about what’s good diet and exercising plans and how to portion control and ’cause I know a lot of people don’t know that when they become diabetic. They don’t, they don’t know that. Because they’re used to kind of eating whatever they want*.*”*

Patient 1 mentioned: *“Maybe like suggestions for different things like they want me to eat three meals and three snacks and there is no way I can keep all that down half the time, so maybe like suggestions or like what could be a snack and things like that and meal suggestions ’cause you don’t want me eating carbs and go easy on the sugar*.*”* They also mentioned: “*They tell me what they want them to be, like they want it to be 125 after a meal and I can’t get 125 to save my life. So, I’m just thinking like how am I going to get to this level. So they have me at like 10, 10 units of insulin; I think using the app would be easier with that part ’cause if like that 10 units of insulin is not enough, it needs to increase. I think being able to keep track of that in the app would be helpful too. Like hey if this meal has this much carbs, take this much insulin, ’cause I know it’s not like a sliding scale like it was before. So now they’re like you need to take this set amount, so I think just having some more reference material for the sugars in general. Like how the insulin makes you feel, what you should do if it’s low, things like that, you know, because sometimes what, and when it’s low I don’t know what I, what I should drink, what I should eat because before if it fell low I would get some juice or something, but now I’m scared the juice is going to raise it too high, you know what I mean? So that would be kind of helpful too*.*”*

### Provider-reported Needs and Preferences in a Dashboard Application

Providers indicated a general set of expectations for a dashboard application including features such as dynamic and actionable data, a method to capture unmet social needs, and enhanced patient-provider communication.

#### Data that is Dynamic and Actionable

Most physicians expressed the need for electronic and standardized means to capture blood glucose data. Typically, providers receive blood glucose values through email and paper logs brought to a clinical encounter. Provider 2 acknowledged the value of having a more comprehensive story about their patient at their fingertips: *“Estimated fetal weight, the gestational age of the patient, the dosing of the insulin or metformin they’re on, if I could have all that information, I could do a lot of this work within the app*.*”* Providers indicated the need for an application that would account for different preferences. One provider expressed the need to have the ability to construct graphs and visualize data in a graphical form.

Provider 7 shared: *“I think an ideal dashboard portal would allow for easy, timely access to blood sugar data, it would be in a mechanism that’s … more dynamic and that can be reviewed, where you can then calculate data, graph data and look at over time on, so it would be more, it would be data that is more easy to encounter and more easy to manipulate and easier to review*.*”*

Another provider, Provider 3, noted the importance of a user-friendly application: *“Right, um, obviously there, there are a lot of systems now available for displaying, for instance, blood glucose levels to providers. I’ll start there, and I’ll say that in my opinion, most of them do not represent user friendly displays. And an incredible amount of data is collected in some of these systems. Yet it’s not user friendly to the practitioner, especially in pregnancy, where decisions are made week to week, if not even more frequently with regard to changes in insulin dosage*.*”*

#### Unmet Social Needs

Providers expressed that there was no standard procedure to capture unmet social needs information. Provider 3 noted: *“You know where there are, is some, albeit skeletal social work support and what have you, but I’m not sure that that’s uniform with them … I think that when that becomes very apparent is when an individual woman indicates that you know, resources are an issue for her in terms of doing glucose testing in terms of obtaining medication including insulin and then that comes to a head pretty quickly, and then we have to do some problem solving*.*”*

Further, providers indicated that it would be helpful to view information collected by patients for any unmet social needs. As mentioned by Provider 7: “*Often patients are the best, I mean, they know if they’re exposed to smoke or if they have access to healthy food or you know what kind of safe neighborhood or lack thereof, they live in, or if they’re able to get exercise and whatnot. So rather than having the providers sit there and ask these questions and put the data in I wonder at some level if this can be data that the patient themselves directly enters via an app and then that populated in the dashboard and then the provider is simply reviewing that data or embellishing it or correcting it in the sense that you know they’re not entering it all, and the patient has done the hard work, because the patient knows that stuff best and then the provider is simply reviewing that data, but that it gets entered in a systematic way that you can see it*.*”*

Provider 4 indicated the value of an application that can recommend resources to patients: *“In our clinic and another facility we have one or two social workers and we’re seeing hundreds of patients. You know, every day we’re seeing enormous numbers of patients. In our diabetes clinic on Tuesday we have you know 25 to 35 prenatal visits for our patients with diabetes … the real challenge is, is access to our social workers and that the app may be able to provide a more accessible resource for our patients if they can look at the app and say where can I get healthy food, where can I obtain my medical supplies, how do I arrange transportation to the clinic*.*”*

#### Enhance Patient-Provider Communication

Providers underscored the need to use data from the dashboard to better communicate with patients about their diabetes and pregnancy. Provider 4 noted: “*Having a way to stay in direct communication, making it as easy as possible for the patient in terms of communicating with the team accessing information about her diabetes management, glucose control, diet would really, I think, be helpful. The more the patient understands the challenges, the better partnership can be established*.”

Provider 6 highlighted the tension between knowing enough about the whole patient in contrast to their diabetes management: “*if we were taking care of the whole patient, we would need more nurses on our diabetes team. Because that would take much more time and you know take our focus away from diabetes and diabetes care and blood sugar management. Probably when I took this job it was hard for me to limit my focus to just diabetes because I had been looking at the whole picture for so long, but I kind of finally learned to hone in, but that doesn’t mean, you don’t ignore things when somebody brings something up, you just have to redirect them*.”

### Define

Upon review of the transcripts, our team identified problems and possibilities faced by our patient population. Three study members used sticky notes to operationalize important concepts onto the Miro board following pre-defined color codes (i.e., orange for keywords, yellow for general summaries/insights, blue for ideas for prototyping, pink for barriers, azure for manual processes, and violet for analysis/decisions) (see appendix figures S1-S4). The study members worked collaboratively to manually sort notes into clusters of similar functional areas (e.g., communication, glucose management). This was conducted for both the patient mHealth and provider dashboard applications.

Study team members further synthesized information into high-level user specification maps (Figure 2) to translate the user requirements into an actionable development plan, highlighting the essential functions and their subthemes for each application. This process also facilitated forming the structure on how the mHealth and dashboard applications would achieve information exchange.

**Figure 2.**
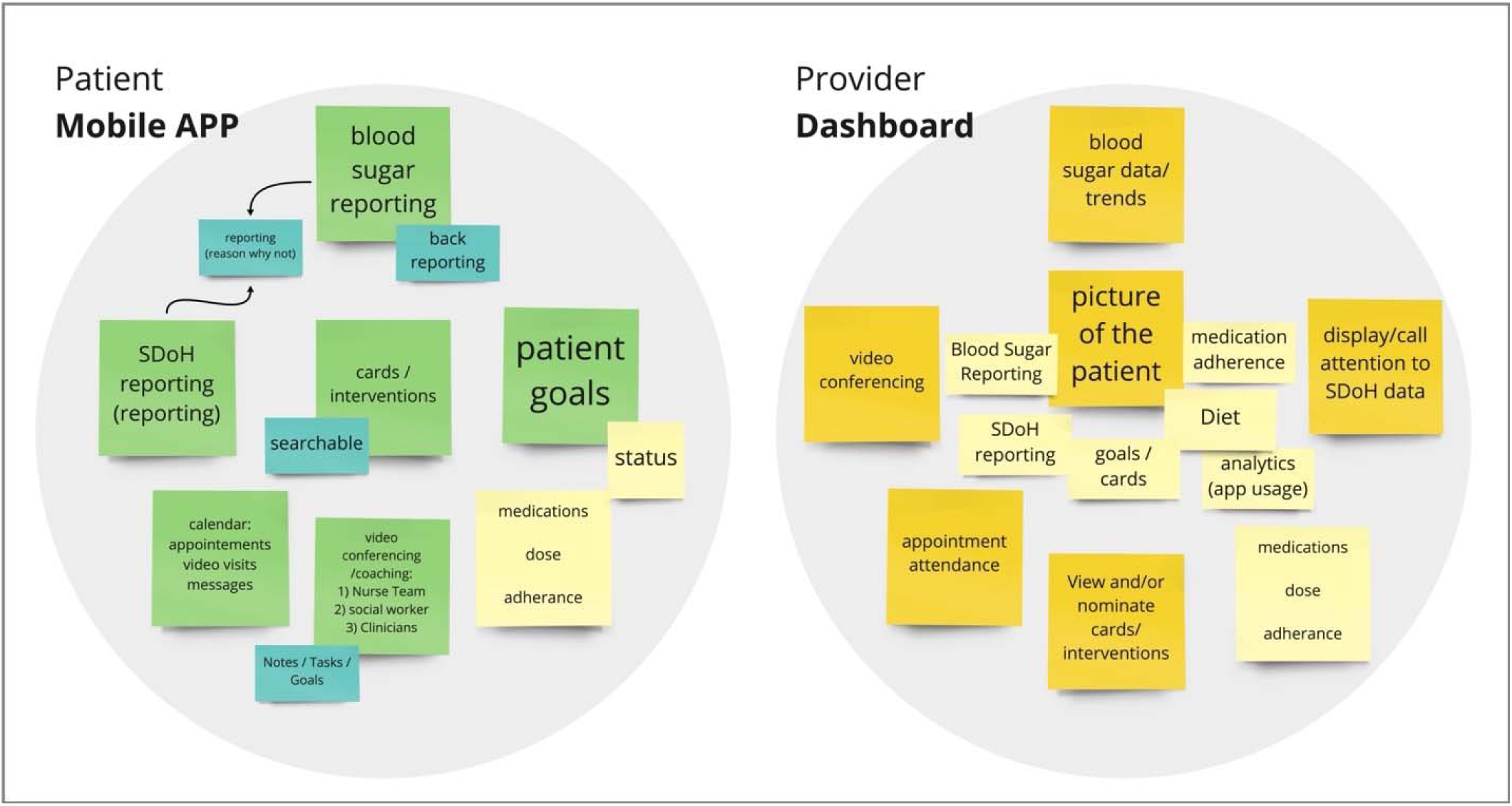
Initial user specifications for mHealth and dashboard applications

The mHealth application centered on the reporting and management of blood glucose values and social needs. Customized patient goals and care pathway cards (clinical and social needs-oriented activities that would help patients achieve their specific goals) were included as primary features, along with other essential elements such as a calendar, video conferencing with the provider team, and medication information. The dashboard focused on showing the trend of a patient’s glycemic control, social needs information, and other critical vitals (e.g., gestational age, other medications) that would present a 360-degree view of the patient. Providers would also use the dashboard to monitor patient activity, actively communicate with them, and tailor their care pathways based on the feedback gathered about a patient from the system.

### Phase II

#### Develop

We continued further analysis of the interview data using swim lane diagrams. A swim lane diagram represents how different roles perform their tasks, take on their responsibilities, and interact with others along a timeline, which generates a holistic view of the system to help identify gaps and discover intervention opportunities. We used the diagram to map the current clinical workflow based on the journey of a Medicaid-enrolled patient with T2D during pregnancy (Figure 3) and developed an improved clinical workflow with the engagement of the applications (Figure 4).

**Figure 3.**
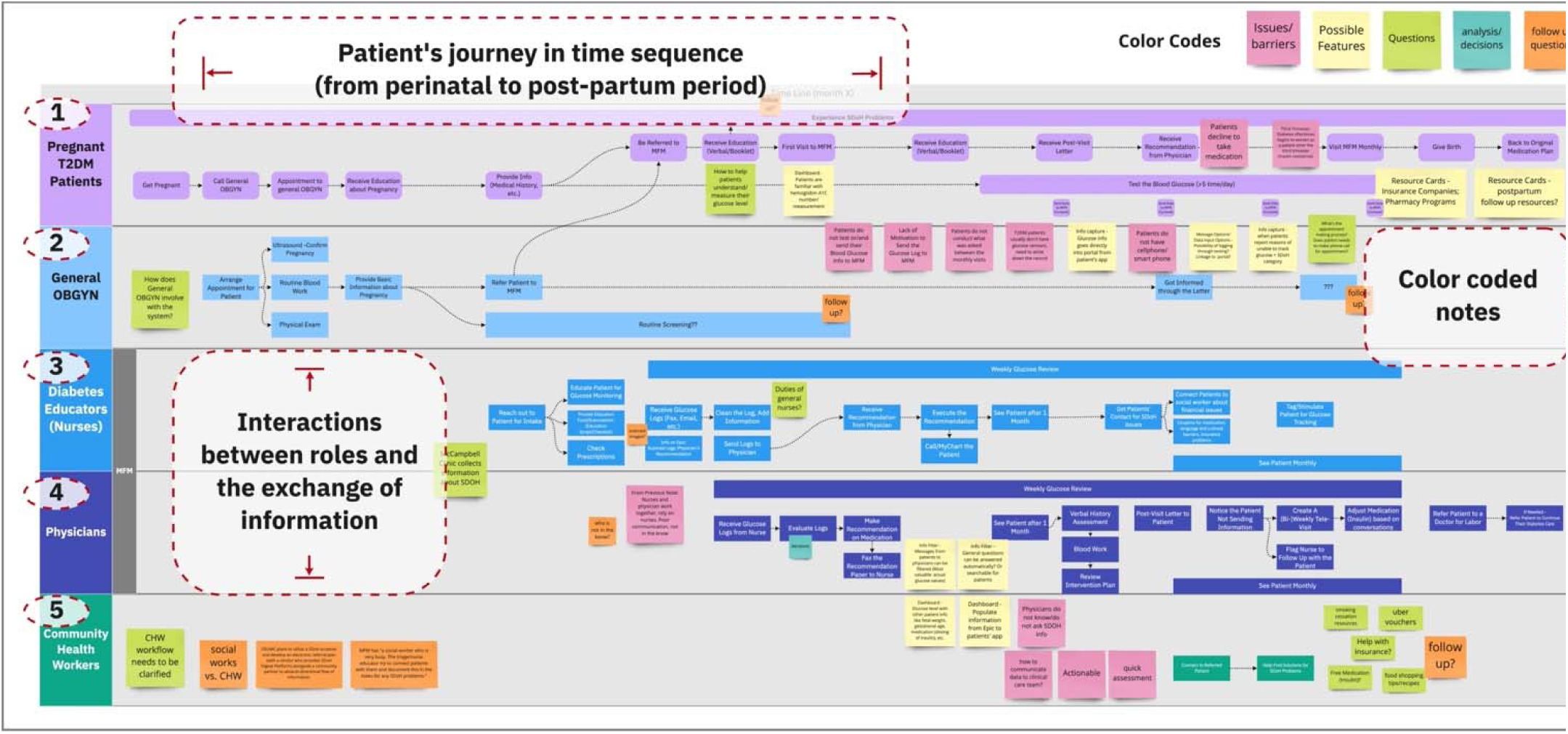
Swim lane of “as-is” clinical workflow (figured scaled to illustrate complexity of the system)

**Figure 4.**
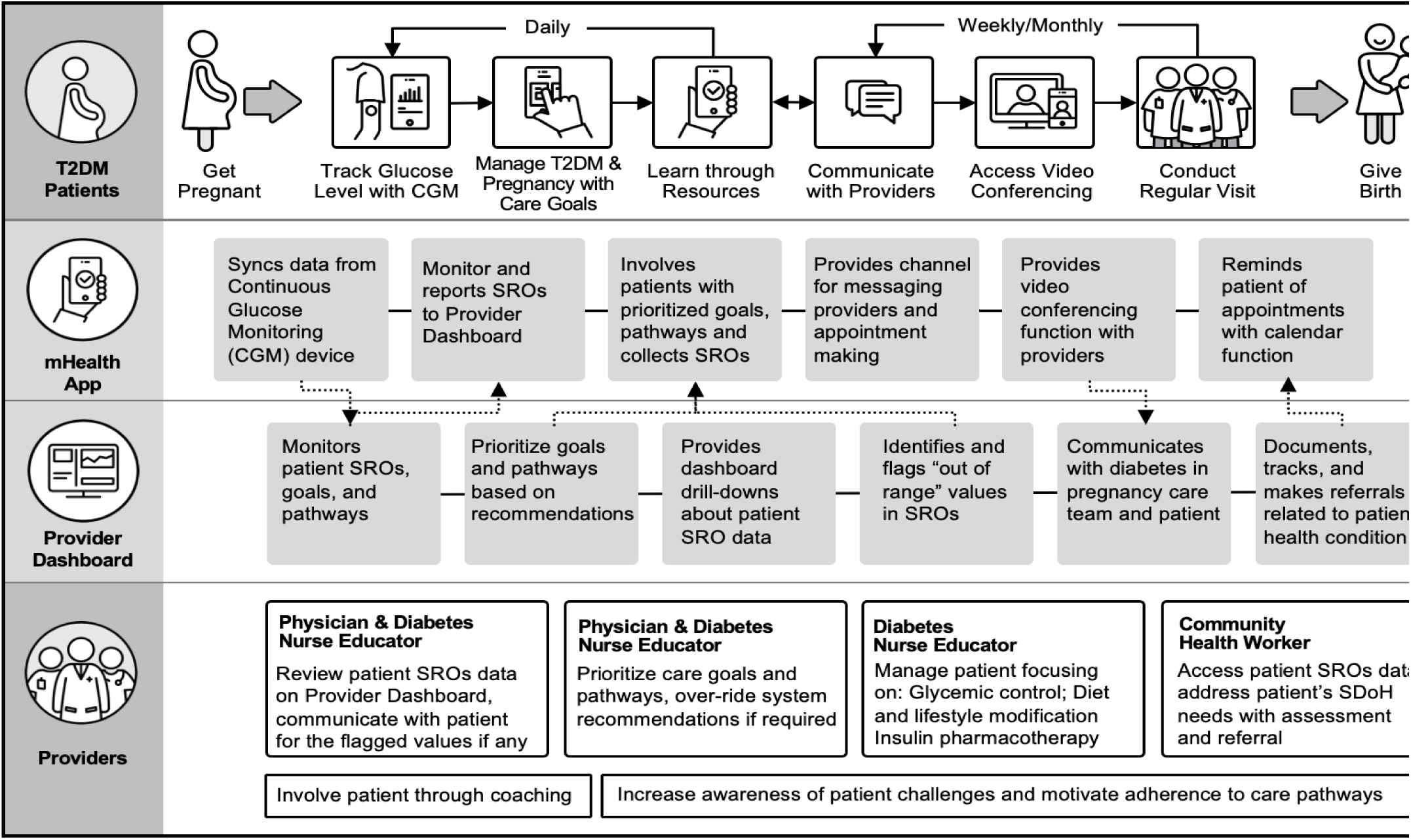
Swim lane of “to-be” clinical workflow with ACHIEVE tools

In the “as-is” clinical workflow (Figure 3), five roles (1-Pregnant T2D Patients, 2-OB/GYN or MFM provider, 3-Diabetes Nurse Educator, 4-Physician, 5-Community Health Worker) are presented in lanes with distinguished colors, followed by the actions taken from a patient’s perinatal period through their post-partum period. The arrows and lines indicate the flow of interaction. We used color-coded notes to capture findings in the workflow, which include issues/barriers, possible features, questions, analysis/decision points, and follow-up questions. During this activity, a variety of gaps were revealed. Major ones included: 1) the patient’s adherence to their goals, care plan, and medication regimen; 2) the communication and information exchange between the patient and the provider, as well as among members of the care team; and 3) the need to address barriers to social needs occurring throughout pregnancy.

Based on the study results from Phase I and our additional analyses, our design team designed high-fidelity prototypes for the mHealth application and dashboard. With features reflecting patient and provider generated requirements with the goal of achieving relevance through a useable, useful, and desirable product [63]. Figure 5 displays the mHealth application prototype with five different pages that cover critical domains identified for the application. Figure 6 displays the provider dashboard prototype with a patient page displayed. Both prototypes were used for our usability testing.

**Figure 5.**
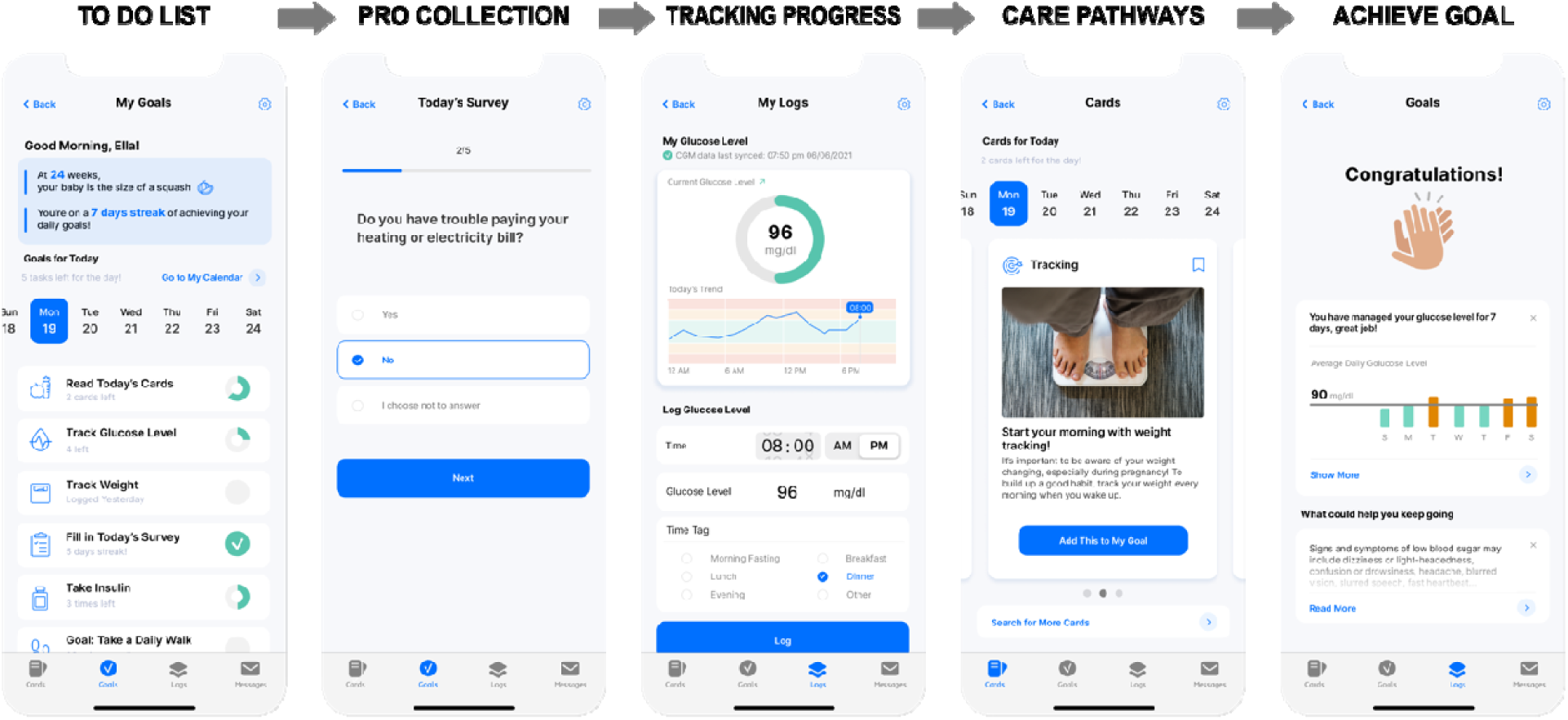
mHealth application prototype. The application allows a patient to view a to-do list that offers an overview of their tasks for the day; report patient reported outcomes; track their glucose levels; manage and learn about their care pathways; and track their goal achievements over time (names are not real).

**Figure 6.**
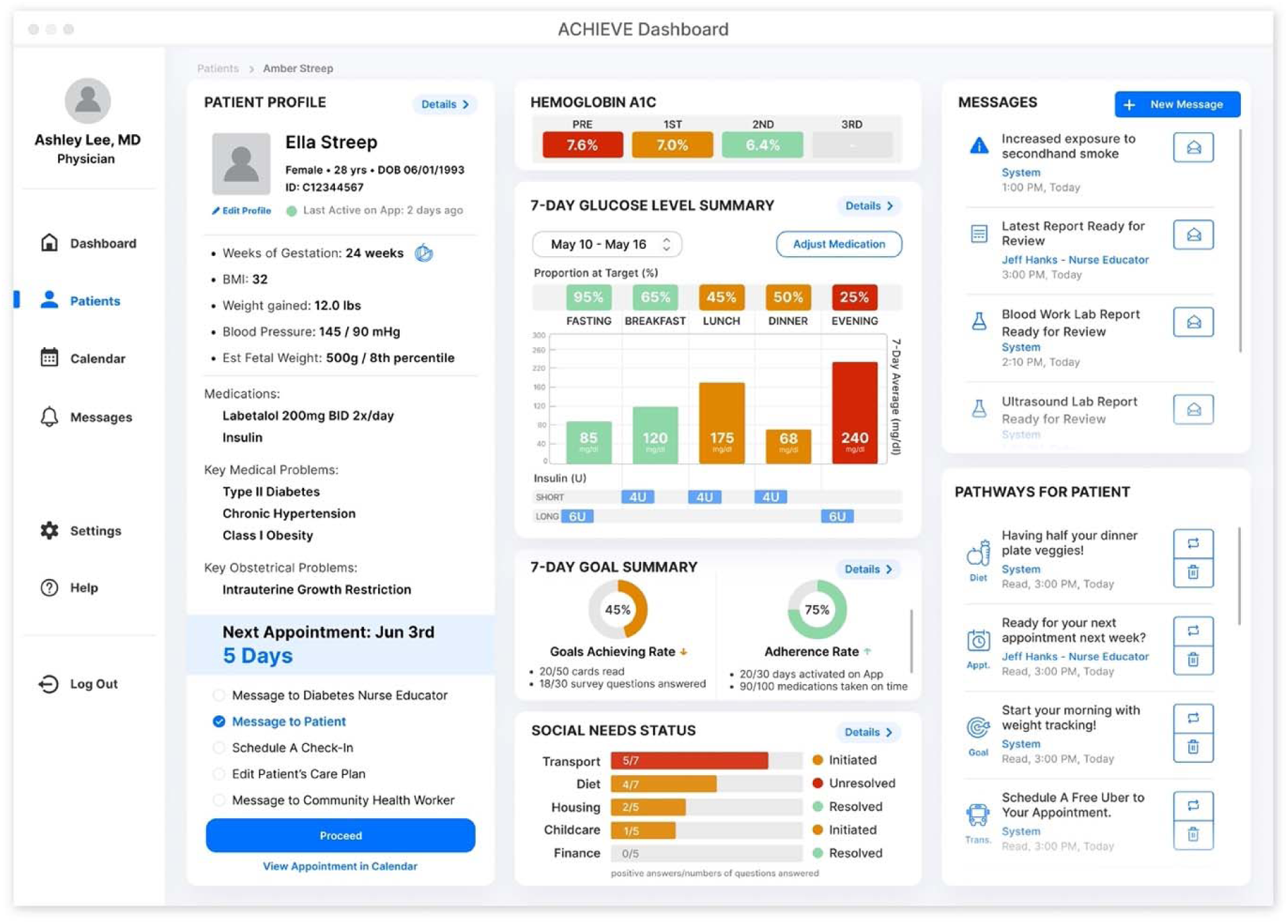
Dashboard application prototype. The dashboard presents patient profiles, scheduling information, and options to communicate with the patient or provider; track A1c, proportion of time in the target range for glucose level, their goal achievement, and social needs; and see notification messages about the patient’s health status and care pathway selections for the patient given their specific patient reported outcome responses (names are not real).

### Deliver

We conducted usability assessments of the mHealth and dashboard applications with patient and provider participants, respectively. We synthesized and categorized feedback from participants and their actions into the content, function, and aesthetics domains based on the application tested (Tables 3 and 4).

**Table 3:** Patient feedback and recommendations for the mHealth prototype

| Themes | Problem | Rationale |
| --- | --- | --- |
| Content | More details about the achievement of goals | Chronicling the achievement of goals and providing additional details about how much more is required for each goal would help patients stay motivated. |
| Function | Initial interaction with the application should have help tool tips to guide user | Patients were lost with how to input the blood glucose levels for specific times, set up appointments, or find recommendations the first time they attempted. |
|  | Include a reminder on the calendar | Patients with busy schedules requested reminders for important events related to their diabetes and pregnancy care. |
|  | Add an alert to indicate when diabetes prescriptions are ready | Patients would like to know when an order is filled for needles and medications so they do not go to pick them up only to find out the pharmacy did not receive the order yet. |
|  | Provide access to a 24-hour call line | Provide patients with the ability to get immediate feedback and advice about problems. |
|  | Add help and educational tabs | It would take time to get used to the application and a help feature would support the learning process. An educational application would help with preventing patients from using Google to obtain information, which could be misinformation or inaccurate. |
| Aesthetics | Relabel goals screen to home screen | The goals page is the home screen, and it has a lot of information other than goals. It should say home because it is greeting the patient, and it has their calendar and other options. |
|  | Use different colors to indicate status of goal completion | The use of blue alone is inadequate and is too similar to the blue highlight on the calendar. |

**Table 4:** Provider feedback and recommendations for the dashboard prototype

| Themes | Problem | Rationale |
| --- | --- | --- |
| Content | Provide median blood glucose numbers and proportions of highs and lows | Averages can be easily distorted; knowing the number of high and low events is valuable to providers. |
|  | Include information about patient's weight | Diabetes dosage requires knowing the body mass of the patient. |
|  | Clarify legend for social needs, explain each dimension, and add more dimensions | The current visualization is too complicated and requires additional clarification about each social need (transportation, financial, etc.) and the incorporation of other dimensions such as housing and childcare needs. |
|  | Include information about changes in diabetes medication dosage and the brands used | Physicians are less likely to adjust dosage if a recent dose change occurred, and knowing the brand would help with ensuring adherence to specific medications. |
|  | Provide additional clarification about patient glucose logs with notes by patients | Patients' notes with their logs would allow providers to know specifics about their daily activities for context for unusual blood glucose readings. |
| Function | Include an alert for consistently high/low blood glucose levels | Providers would like the convenience of quickly identifying problems with patients' ability to manage their blood glucose. |
|  | Allow for default choice of blood glucose summary or detailed log and allow for drill downs | Providers have different preferences for either a summary or varying levels of detail (days, weeks, overnight numbers) of the blood glucose logs as the default view. |
|  | Add a task list with specified roles | A task list would allow care team members to be aware of which member would be responsible for addressing a specific patient problem. |
|  | Add checklist for completed educational resources | Diabetes educators can monitor and ensure that patients have reviewed and understood specific educational components assigned to them. |
|  | Add dosage of diabetes medication | Providers would be able to make better decisions for prescriptions for patients if they knew the dosage of all the diabetes medication patients took. |
|  | Add a log to track patient meals | Allow providers to be able to educate patients about the connection between what they are eating, especially carbohydrates, and their blood glucose levels and make recommendations for them. |
|  | Add allergy to insulin information | Knowing which insulin the patient is allergic to would allow the provider to make safer prescriptions through the pharmacy and avoid adverse events. |
| Aesthetics | Use a different color for the bar graph to indicate a low blood glucose level | The colors for the bar graphs were categorized by meal and could make it harder for a provider to quickly identify low values. |
|  | Create a visual hierarchy or minimized view with the option for drill down in the left most column to create space for other critical components | Physicians may overlook the left column and not look at it frequently because of the patient clinical history information presented. |
|  | Rename cards to resources | Patients did not find the label cards to resonate with them and preferred resources. |

#### Task load survey

Workload measurements such as the TLX provide a general sense of how tasks related to the use of a tool are experienced. Overall, TLX unweighted scores reported were 33 (low scores indicate less cognitive load) for both applications, which translated to the 20th percentile of a national review of studies that used the TLX across multiple technologies (overall mean score=42 and health care median score=45). For the mHealth application, this score was driven primarily by relatively low temporal and frustration scores; patients expressed some concerns about the performance demands (possibly due to the lack of initial training to input data). The TLX score for the dashboard was driven primarily by low physical and temporal scores, with equally moderate concerns raised about mental and performance demands. Reweighted scores for both TLX measures indicated improvements, with a higher difference for the dashboard. The reweighted measure reallocated a total weighting of 15 points across four of the five domains (mental=4; performance=3; effort=2; frustration=3; physical=0) as recommended by Hertzum and colleagues based on the study of similar technologies reported in the literature [64].

**Table 4:** Unweighted Patient and Provider TLX scores

|  | **Patient mHealth application Score (SD)** | **Provider dashboard application Score (SD)** |
| --- | --- | --- |
| Overall | 33 ($\pm 9.3$) | 33 ($\pm 4.4$) |
| Domain |  |  |
| Mental | 33 ($\pm 21$) | 50 ($\pm 10$) |
| Physical | 27 ($\pm 29$) | 13 ($\pm 6$) |
| Temporal | 17 ($\pm 12$) | 20 ($\pm 10$) |
| Performance | 53 ($\pm 20$) | 53 ($\pm 15$) |
| Effort | 37 ($\pm 6$) | 37 ($\pm 6$) |
| Frustration | 27 ($\pm 6$) | 27 ($\pm 21$) |

## Discussion

Stand-alone mHealth applications for diabetes management in pregnant individuals exist, but only a few are comprehensive and integrated, and none are tailored to meet the needs of those on Medicaid [21]. We leveraged findings from existing studies to design an integrated approach for Medicaid-enrolled individuals with T2D during pregnancy that provides a comprehensive, tailored, and team-based solution addressing clinical management and social needs using themes identified through prior research, behavioral theory, and our qualitative work. Our intervention brings together multiple technologies in an integrative framework to gain a 360-degree view of a T2D patient’s pregnancy experience more effectively and be responsive to that information during the pregnancy trajectory regarding clinical, social, and supportive care.

### Principal Findings

We identified three themes that exemplified pregnancy experience among Medicaid enrolled individuals with T2D: *managing exhaustion, adherence to a new regimen, and preparing for pregnancy*. Exhaustion and fatigue are among the commonly reported feelings during pregnancy that may exacerbate a chronic comorbid condition, such as diabetes [65]. During pregnancy with diabetes, specific lifestyle adjustments are recommended over a short period of time. [7] Individuals with T2D who were previously on oral pharmacotherapy or diet-based therapy now generally require insulin to achieve glucose control using strict parameters, including upwards of four daily FSBG [66,67], and patients are challenged to learn to regulate both the quantity and quality of carbohydrate consumption while simultaneously adopting new medication use and exercise behaviors. The strict medication and dietary requirements and increased home-based activities such as glucose monitoring and reporting escalate the burden of disease management contributing to pregnancy fatigue. Moreover, multiparous women taking care of older children may face additional barriers that compound this preexisting exhaustion.

Diabetes in pregnancy requires economic and social stability. T2D management in pregnancy is expensive, with >$7,000 in excess pregnancy-related costs, and individuals often report financial planning as part of diabetes management strategy [68]. Likewise, social support is equally important for good quality of life during pregnancy and requires planning in the earlier stages. The themes we identified align with previous finding. The associated disease novelty, especially among nulliparous women living with T2D, and dynamic physiological and clinical changes require preparation for upcoming lifestyle demands [69,70].

Participants expressed needs and preferences in an mHealth application that were represented by five themes: *electronically captured and managed information, access to support systems, use of diabetes technology, help through problem solving, and personalized recommendations*. The most desired feature in an mHealth application was the ability to electronically capture blood glucose data. Pregnant individuals with diabetes often find it cumbersome to manually record, save, and communicate their glucose values with their care team providers. An electronic platform with the ability to capture and communicate blood glucose values in a meaningful manner emerged as a desired feature by both patients and providers. As opposed to manual data entry, an mHealth application provides patients with the opportunities to experience streamlined and routinized data collection that requires less effort on their part. For example, use of a CGM device that syncs with an mHealth application with a user-friendly interface may promote long-term use of the application and device [71].

### User Needs and Preferences for T2D Management during Pregnancy

The interrelated themes we identified from patients and providers revealed specific end-user specifications that aligned with the theoretical underpinnings of SCT. SCT posited that successful performance of a behavior depends on an individual’s behavioral capability as well as cognitive and environmental influences on behavior via three domains: 1) skills; 2) knowledge and beliefs; and 3) self-efficacy [44]. The themes identified from our qualitative work highlight the importance of these three domains specific to individuals with T2D during pregnancy. Individuals require educational information that clearly explains how behaviors and specific clinical and social needs pathways will help them achieve the primary outcome of glycemic control (knowledge and beliefs). Patients need skills and coaching to engage with the diabetes care team, which can be accomplished by collecting and synthesizing detailed information about the pregnant individual from the mHealth application to the provider dashboard to better communicate with the diabetes care team (skills). Team-based coaching based on CGM output, personalized information, and care pathways may help the pregnant individual learn more about T2D in pregnancy and better adhere to their T2D care regimen. Closing the loop (e.g., having a community health worker document the individual’s successful securing of healthy food through a community food pantry) could ensure that an individual’s new skills yield meaningful outcomes and enhance their confidence (self-efficacy).

### Design Requirements and Refinements Needed to Meet End-User Preferences and Needs

Our research team recognized the importance of leveraging technology that is highly relevant to our study population and their providers. We utilized applications that did not require high costs to maintain in the long term and were evidenced-based. In our exploration of the evidence base for mHealth and provider dashboard applications tailored to similar populations, we quickly recognized the challenges faced in prior implementations of each application [21]. Specifically, these previous approaches to improving care for Medicaid-enrolled patients with diabetes have been siloed and fragmented [11]. We therefore identified an intervention that would be systems-oriented and addressed the pitfalls associated with the use of each application by itself. We focused more on the gestalt effect of these applications and their ability to collectively transform health care by providing a comprehensive, 360-degree view of the patient using information that was dynamic and closed the loop between the provider and the patient (e.g., the provider will be aware that a patient completed an assigned care pathway).

Our analysis of the “as-is” and the “to-be” workflows identified major gaps: 1) patient adherence to their goals, care plan, and medication regiment; 2) the communication and information exchange between the patient and the provider, as well as among members of the care team; and 3) the need to address barriers to social needs occurring throughout the pregnancy. These were critical to an ideal workflow. Our applications considered not just the clinical and psycho-social requirements for the patient, but also these realities in our clinical setting. We pursued a deeper understanding of the requirements of applications by testing them with patients and providers, and although initial impressions were highly positive (e.g., TLX scores for both applications were in the 20th percentile of national TLX scores across all devices), we obtained notable feedback that will be used to improve the content, function, and aesthetics of our applications before a full deployment. Our favorable scores on the TLX surveys for both our prototypes should also be highlighted because participants did not receive any pre-training to use the applications. This suggests the intuitiveness of our prototypes, especially the interfaces. The implications for such design performance may include reductions in cost and effort associated with training and greater engagement from individuals with lower technology literacy.

### Implications and Recommendations for Future Design and Practice

A key consideration when it comes to the use of digital health tools is the reversal of newly established behaviors. [72,73] Whether our approach – which is supported by prior studies, our own mixed-methods findings, and theory – results in long-term behavioral modifications is not demonstrable in this paper and requires further inquiry. A critical component of such an inquiry would involve gauging the cost-benefit ratio that individuals mentally construct in determining their desire to balance the use of our applications to manage blood glucose levels during their pregnancy. Relatedly, the timing of our intervention is another area of consideration, especially given our focus on pregnancy. Extending the use of our application before pregnancy (in preparation for pregnancy) and postpartum use (especially during the interconception period) could have implications for the successful management of blood glucose beyond the perinatal period.

### Strengths and Limitations

An important limitation to our study is that it is not tied to the evaluation of health outcomes. It reflects realistic perspectives of patients and providers whose behaviors and interactions could have an impact on health outcomes. Our study is also focused on a specific population during pregnancy and does not capture their views before or after pregnancy. Our study involves perspectives of patients and providers who are highly committed and engaged in the use of our applications, which may influence study findings [74]. Nevertheless, our team was able to gather both positive and negative impressions on the final prototypes. More work is required to obtain generalizable perspectives beyond our sample. Our approach to developing applications using a double diamond framework with a participatory design mindset is generalizable and is already utilized in multiple domains beyond health care including government, finance, insurance, and other general consumer markets.

## Conclusions

Digital health tools such as mHealth and dashboard applications have the ability to transform health care among Medicaid-enrolled patients with T2D during pregnancy, with the goal of managing their blood glucose levels, which is a precursor to experiencing a successful pregnancy and birth. Distilling patient and provider needs and preferences, then using them, along with prior studies and theory, to develop applications, holds great potential in tackling complicated health care issues. Engaging individuals such as those from our study population presented a unique set of challenges, and it was apparent to our team that system relevance throughout pregnancy was a critical success factor for our end-users relative to all others, regardless of efficacy. The methods described in our study can be used as a template for future design considerations specific to the development of digital health interventions, including those focused on understudied populations.

## Data Availability

All data produced in the present work are contained in the manuscript.

