## Supplementary figures for "Developing and testing an integrated patient mHealth and provider dashboard application system for type 2 diabetes management among Medicaid-enrolled pregnant individuals based on a user-centered approach: Mixed-methods study"

Figures S1 to S4 illustrate the process used by the study members to ideate and prioritize system requirements based on the literature review and interviews.

**Figure S1.** Cross-sectional screen shot of initial themes identified for dashboard application from providers’ perspectives

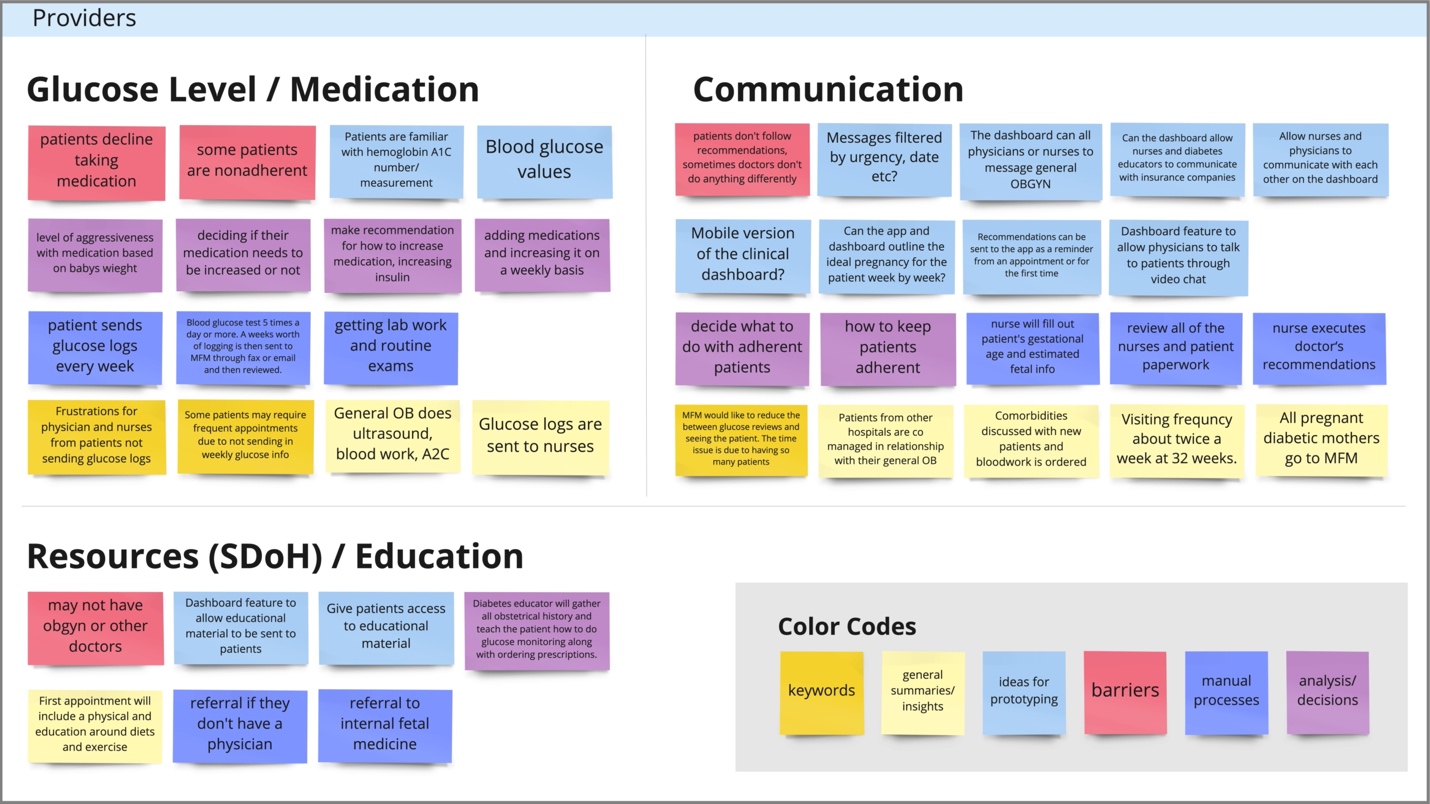

**Figure S2.** Cross-sectional screen shot of initial themes identified for mHealth application from patients’ perspectives – Cluster 1. Data Tracking

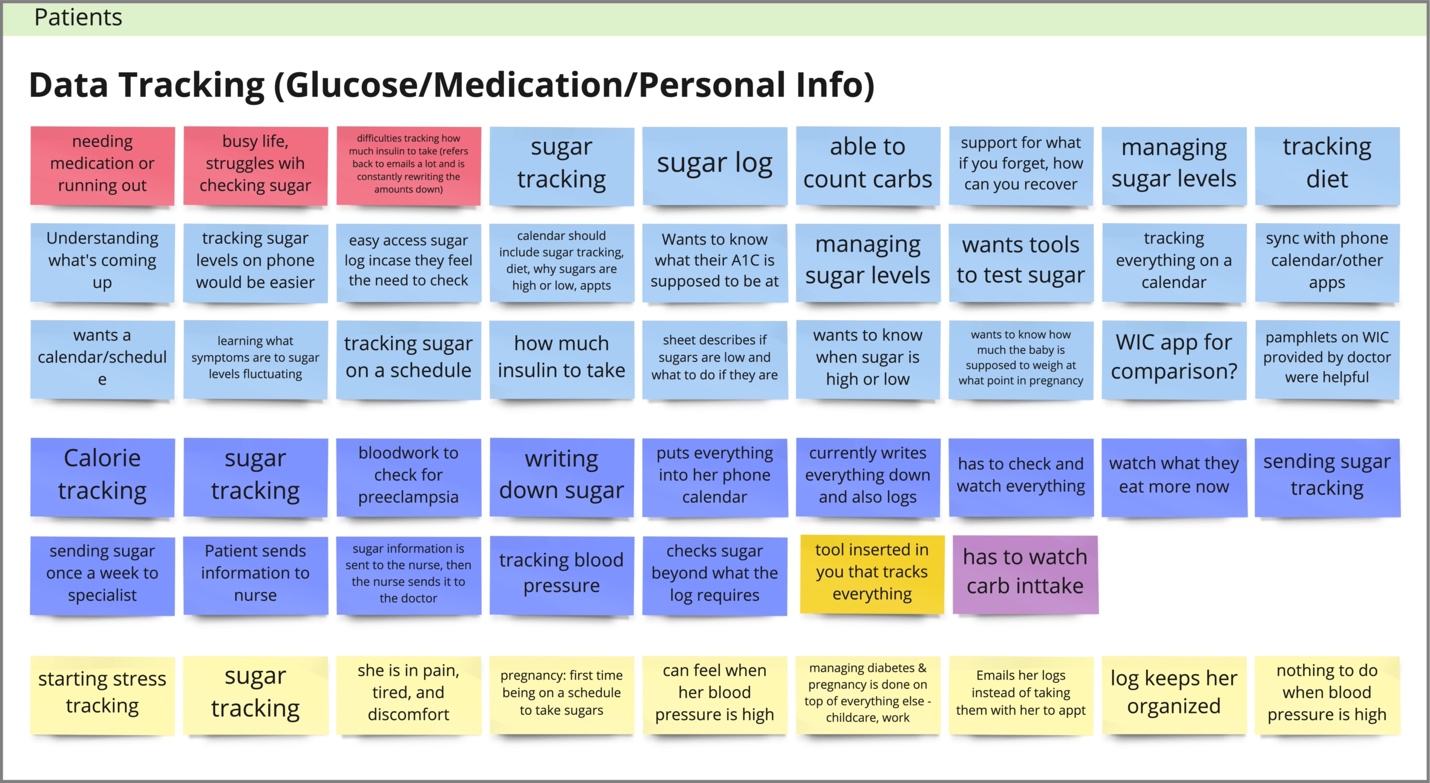

**Figure S3.** Cross-sectional screen shot of initial themes identified for mHealth application from patients’ perspectives – Cluster 2. Communication

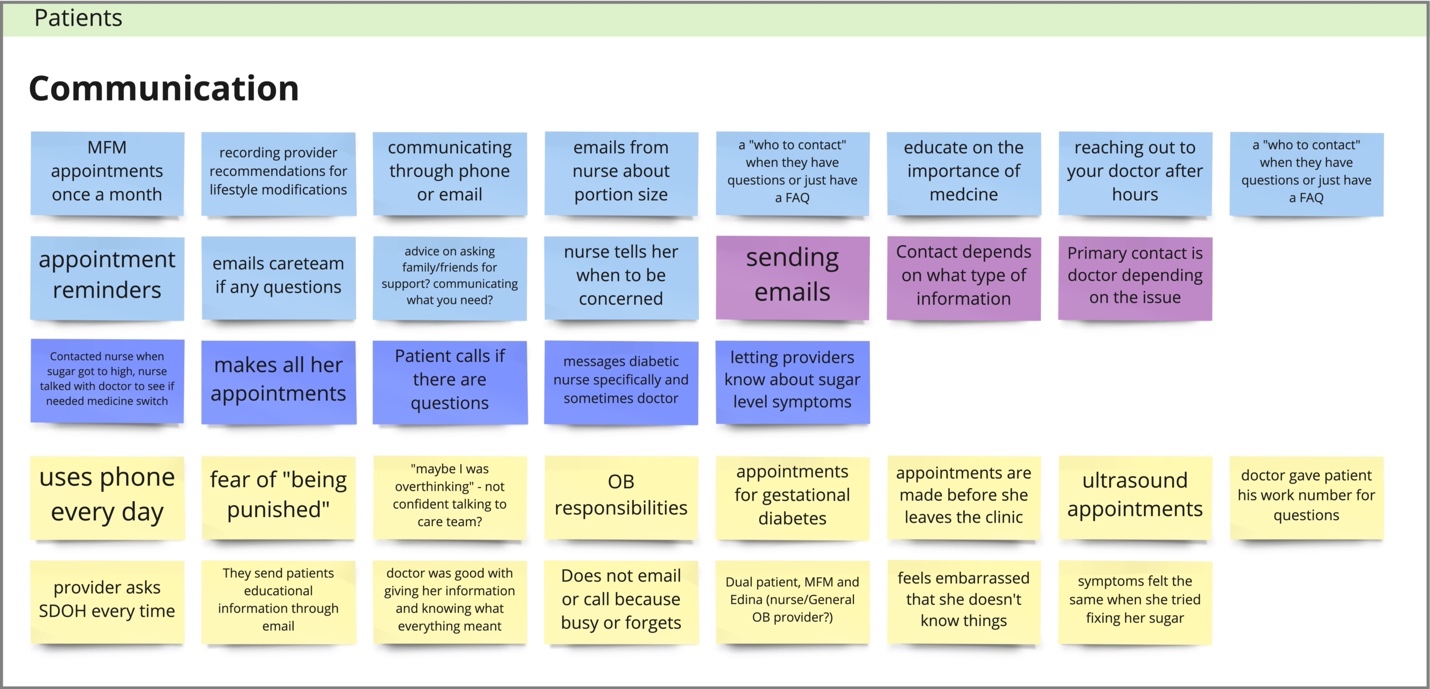

**Figure S4.** Cross-sectional screen shot of initial themes identified for mHealth application from patients’ perspectives – Cluster 3. Resources

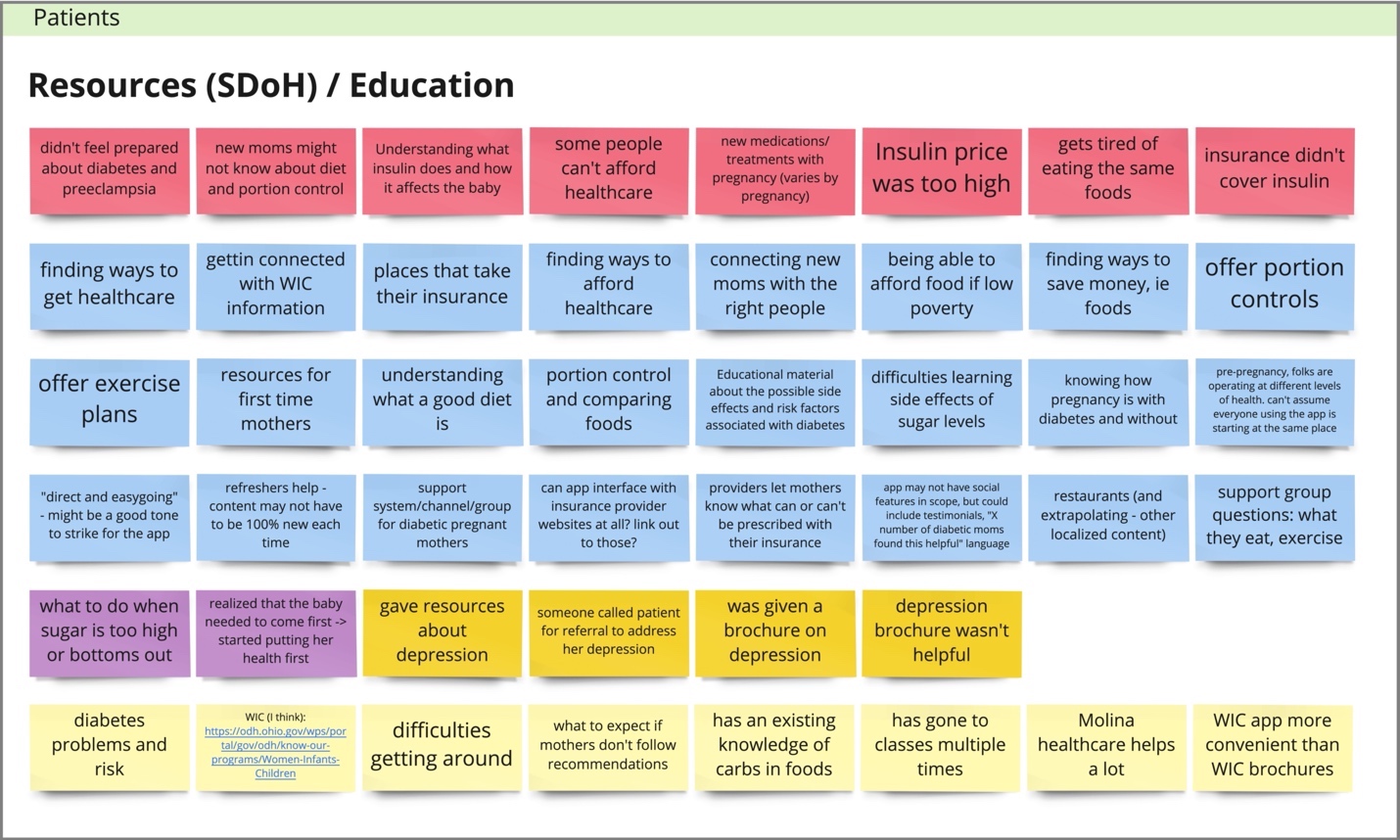
